# Toward Generalizable and Clinically Useful Prediction of Occupational Functioning in Schizophrenia Using a Structure-Informed Representation of Cognition

**DOI:** 10.64898/2026.06.25.26356524

**Authors:** Chen Chen

## Abstract

Predicting real-world functioning remains a clinical priority in schizophrenia (SCZ), but existing prediction models face methodological limitations and lack established clinical utility. Cognition is a commonly used predictor and how it is represented may affect predictive performance. The Normative Latent Cognitive Structure (N-LCS) approach provides a structure-informed representation that may address limitations of conventional domain-level scores. Data from the COBRE cohort (163 SCZ, 180 healthy controls [HC]) were used to develop and internally validate ridge regression models for occupational (OF) and social (SF) functioning using N-LCS deviation metrics alongside a priori selected demographic and clinical predictors within a fully nested, optimism-corrected bootstrap framework. The OF model achieved an optimism-corrected AUC of 0.73 and balanced accuracy of 0.71, with near-ideal calibration and net benefit across nearly the full range of threshold probabilities (0-0.99) on decision curve analysis. The SF model showed modest performance (C-index = 0.66, weighted kappa = 0.27). A reduced OF model was externally validated in an independent cohort (NUSDAST; 166 SCZ, 156 HC), maintaining discrimination (AUC = 0.70) and showing improved balanced accuracy (0.56 to 0.69) after recalibration. Compared with the conventional score-based model, the N-LCS OF model achieved comparable predictive performance with fewer predictors and better calibration. Overall, these findings support the potential of the OF model as a generalizable tool for predicting occupational functioning in SCZ.

## Introduction

Schizophrenia (SCZ) is associated with marked impairments in real-world functioning. Employment rates remain below 20% [1], and deficits in social participation and community integration persist even among patients with well-controlled psychotic symptoms [2]. Such difficulties impose a long-term burden on patients and society and remain poorly addressed by current treatments [3].

Accurate prediction of functional outcomes could facilitate early identification of patients who may benefit from rehabilitation and vocational support, enabling more individualized intervention planning. However, despite rapid growth of psychiatric prediction modeling, recent systematic reviews have highlighted methodological limitations in many published models, including bias and overfitting due to suboptimal model-development and validation practices, and insufficient attention to calibration, clinical utility, and independent external validation [4, 5]. As a result, reported model performances are often optimistic and unlikely to hold in new patients [6], limiting confidence in their clinical applicability.

In addition to methodological considerations, how predictors are represented may also influence predictive performance. Cognitive measures are among the most frequently used predictors of functional outcome in SCZ [7], but they are often represented as domain-level scores derived from standardized neuropsychological batteries, which may not fully capture the cognitive heterogeneity observed in SCZ [8]. Differences across assessment batteries further limit the generalizability of models developed in one clinical setting to others [9], and interpretation of these scores is complicated by task impurity, as individual tasks often engage multiple cognitive processes [10]. These limitations may partly explain the inconsistent performance of cognition-based prediction models across studies and cohorts, highlighting the need for alternative representations of cognition to improve prediction of real-world functioning.

One such approach is the Normative Latent Cognitive Structure (N-LCS), which provides an individualized, structure-informed representation of cognition. N-LCS has demonstrated structural consistency across independent cohorts assessed with different cognitive batteries, supporting its potential as a transferable cognitive representation for prediction modeling across cohorts, and its deviation metrics have also shown symptom associations beyond those detected by score-based measures [8]. Whether these advantages translate into improved prediction of real-world functional outcomes remains to be determined.

Building on this evidence, the present study had three aims. The primary aim was to develop and internally validate prediction models for real-world functional outcomes in SCZ using N-LCS- derived metrics, following current recommendations for rigorous clinical prediction model development [4]. The best-performing model was then externally validated in an independent cohort to evaluate its generalizability. Finally, the performance of the N-LCS models was compared with that of conventional score-based models using the same predictor framework.

## Methods

### Data

The model development dataset was obtained from two SCZ cohorts funded by the Center of Biomedical Research Excellence (COBRE) program [11, 12], comprising 163 patients with SCZ and 180 healthy controls (HC) after merging datasets. Prior to merging, the two cohorts were compared on all demographic, cognitive, and (within the SCZ group) clinical/functional variables used in subsequent analyses, to assess comparability; no significant differences were observed after false discovery rate (FDR) correction (all p > 0.92). Data were downloaded via the Collaborative Informatics and Neuroimaging Suite Data Exchange (COINS) tool (http://coins.mrn.org/dx). Participants were recruited at the University of New Mexico. Patients were clinically stable at the time of assessment, with controlled symptoms and antipsychotic medication for a minimum of 3 weeks.

An independent cohort obtained from the Northwestern University Schizophrenia Data and Software Tool (NUSDAST) was used for external validation of the developed models [13]. Data were downloaded from XNAT Central (https://central.xnat.org/). The NUSDAST cohort included 166 patients with SCZ and 156 HC. Patients were recruited from local inpatient and outpatient treatment facilities and were stabilized on antipsychotic medication for at least 2 weeks prior to assessment [14].

All data were fully anonymized, and the present analyses did not require additional ethical approval.

### Cognitive measures

In the COBRE cohort, cognitive function was evaluated with the MATRICS Consensus Cognitive Battery (MCCB) [9, 15]. Domain T-scores for processing speed, attention, working memory, and reasoning were used in the analyses, consistent with the domain selection established in the original N-LCS study [8]. MCCB domain scores were derived from the T-scores of their constituent measures and standardized using the MCCB normative sample [15].

In the NUSDAST cohort, cognitive performance was represented by four domain scores, including crystallized intelligence, working memory, episodic memory, and executive function. Domain scores were calculated based on the constituent neuropsychological tests within each domain and converted into z-scores based on the combined SCZ and HC sample (2-group calculation) [13]. The cognitive measures used in NUSDAST differed from those in MCCB.

### Clinical measures

In the COBRE cohort, negative symptoms were quantified as the sum of the corresponding items from the Positive and Negative Syndrome Scale (PANSS) [16]. Patient life circumstances were assessed using Lehman’s Quality of Life Interview (QOLI) [17]. Antipsychotic medication dosage was quantified as total chlorpromazine equivalent dose (CPZ).

### Data preprocessing

Data were screened for missing values and outliers before analysis, and participants with missing data were excluded. Cognitive variables in both cohorts were assessed for skewness and standardized using HC-based z-scores, such that SCZ scores were expressed relative to the HC normative reference. In the COBRE cohort, CPZ was assessed for skewness and log-transformed accordingly.

### N-LCS and its deviation metrics

N-LCS was constructed in each cohort as previously described [8]. Briefly, the N-LCS was derived by applying factor analysis to cognitive variables in the HC group. Parallel analysis [18], Velicer’s Minimum Average Partial (MAP) test [19], and bootstrap Tucker congruence [20] were used to evaluate the resulting one-factor solution. The HC sample defined a cognitive space based on its covariance structure, within which N-LCS served as the normative reference. Cognitive data from all participants were subsequently projected into this space.

Two deviation metrics were derived. Cognitive Deviation Angle (CDA) quantified the angular deviation (cosine angle) between an individual’s cognition and the N-LCS, capturing alterations in the relationships among cognitive domains. For the present study, Cognitive Deviation Magnitude (CDM) was defined as the projection of an individual’s cognition onto the N-LCS, reflecting the shared changes across cognitive domains.

To evaluate whether N-LCS represented a comparable latent cognitive structure across cohorts, structural similarity between the COBRE- and NUSDAST-derived N-LCS was assessed using Tucker congruence [20], while directional alignment was quantified using cosine similarity [21].

### Functional outcomes

In the COBRE cohort, two indicators of real-world functioning were derived from the QOLI: occupational functioning (OF; earned income status) and social functioning (SF; frequency of planned activities with others). These outcomes were selected to capture distinct dimensions of real-world functioning in SCZ, rather than relying on composite global functioning measures [22]. Earned income status was selected as the primary OF indicator because it showed high agreement with employment status (Cohen’s κ = 0.88) [23] and provided a larger analyzable sample. This agreement supports the comparability of these two measures and provides a basis for cross-cohort outcome harmonization, as a directly comparable earned income variable was not available in NUSDAST. For the SF outcome, the original five response categories were collapsed into three ordered levels to reduce sparsity in low-frequency categories.

In the NUSDAST cohort, employment status was used to derive the OF outcome for external validation, consistent with the correspondence between employment status and earned income status observed in the development cohort. As NUSDAST did not include a comparable measure of social functioning, external validation was not performed for the SF outcome.

### Candidate predictors

Given the established role of cognition in predicting functional outcomes in SCZ, CDM and CDA were included as novel representations of cognitive functioning. Age, sex, years of education, CPZ, Wechsler Test of Adult Reading (WTAR), and negative symptoms were selected a priori based on existing evidence and their frequent use in prediction models [7, 24–26]. A self-perceived functioning measure derived from the QOLI (“overall functioning in home, social, school, and work settings at the present time”) was also included, as it may capture information beyond objective cognitive performance [27]. All continuous predictors were standardized prior to model fitting.

For external validation, only predictors available in the NUSDAST cohort and included in the final model were evaluated.

### Model development and internal validation

For each outcome in the COBRE cohort, ridge regression was used to reduce overfitting [4]: ridge logistic regression for the binary outcome (OF), and ridge cumulative-link ordinal logistic regression for the ordinal SF outcome. All non-empty subsets of the candidate predictors were evaluated; for each subset, the ridge penalty was tuned via five-fold cross-validation, and the predictor subset with the highest apparent AUC (OF) or apparent C-index (SF) was retained.

To account for optimism introduced during model development, predictor-set selection, penalty tuning, and (for the binary outcome) threshold determination were fully nested within a bootstrap resampling procedure (B = 200) [4, 28]. Optimism was calculated as the difference between apparent performance in the bootstrap sample and performance of the same model when applied to the original sample and averaged across bootstrap replicates. Optimism-corrected estimates were then obtained by subtracting this mean optimism from the apparent performance of the model selected on the original sample.

For OF, the classification threshold was determined using the Youden index within each model (original and bootstrap) [29], and events-per-variable (EPV) was calculated to assess sample adequacy relative to the number of predictors [30]. Optimism-corrected AUC, balanced accuracy, sensitivity, specificity, calibration intercept/slope [31], and net benefit across a grid of threshold probabilities (decision curve analysis (DCA) [32]) were obtained through this nested bootstrap procedure. For SF, model performance was evaluated using optimism-corrected C-index, log-loss, and weighted kappa.

### External validation

External validation was performed using the NUSDAST cohort. Because self-perceived functioning, included in the final OF model, was unavailable in NUSDAST, a reduced model (OF- RM) was re-fit using ridge regression on the same model development dataset (COBRE) after removing this predictor, with the remaining predictor set fixed and without repeating predictor selection. The OF-RM retained similar optimism-corrected discrimination and balanced accuracy to the OF model (see Table 3) and was therefore used for external validation.

The locked OF-RM, including the final regression coefficients and the Youden index, was applied to the independent NUSDAST cohort without model updating. Because CDM was derived relative to the normative cognitive structure within each cohort, only age and education required standardization using the centering and scaling parameters from COBRE. Model performance was evaluated using AUC, balanced accuracy, sensitivity, specificity, calibration intercept and slope, and DCA. Confidence intervals were estimated using 1,000 non-parametric bootstrap resamples with the locked model and classification threshold held constant.

Model recalibration was subsequently performed by updating the calibration intercept and slope. A new Youden index was then determined from the recalibrated predictions, and model performance was re-evaluated.

### Comparison with score-based models

To evaluate whether N-LCS deviation metrics offered advantages over conventional cognitive representations in the COBRE cohort, a parallel model was developed for the OF outcome using MCCB domain T-scores in place of CDM and CDA, with all other predictors in the retained predictor set unchanged. This model was developed and evaluated using the same nested bootstrap framework described above. As the final model for the SF outcome did not include a cognitive predictor, this comparison could not be performed for SF.

### Statistical analysis

Categorical variables were compared using chi-square tests, and continuous variables were examined with Wilcoxon rank-sum tests, with multiple comparisons adjusted using the FDR correction. Effect sizes were calculated using Cohen’s d. Agreement between earned income status and employment status was assessed using Cohen’s κ. Analyses were conducted using all available data for each variable, resulting in minor variations in sample size across measures. Statistical significance was defined as two-tailed p < 0.05. All analyses were conducted in R (version 4.6.0).

## Results

### Demographic, cognitive, and clinical characteristics

A total of 343 participants were included in the model development cohort (COBRE), comprising 180 HC and 163 individuals with SCZ. Demographic, cognitive, and clinical characteristics are presented in Table 1.

**Table 1.** Demographic, cognitive, and clinical characteristics of participants in COBRE cohort.

| Variable | HC | SCZ | p-value |
| --- | --- | --- | --- |
| Age (years) | 37.3 ± 12.0 | 36.8 ± 13.6 | p = 0.61 |
| Sex (Female, %) | 51 (28.3%) | 25 (15.3%) | p = 0.006** |
| Education (years) | 13.8 ± 1.7 | 12.6 ± 1.8 | p < 0.001** |
| WTAR | 108.2 ± 13.0 | 95.9 ± 16.5 | p < 0.001** |
| Processing speed | 52.7 ± 8.9 | 34.2 ± 12.1 | p < 0.001** |
| Attention | 49.5 ± 9.9 | 35.7 ± 14.2 | p < 0.001** |
| Working memory | 47.7 ± 10.5 | 37.2 ± 13.0 | p < 0.001** |
| Reasoning | 54.9 ± 9.1 | 44.2 ± 10.3 | p < 0.001** |
| CPZ (mg/day) | - | 300 (175-600) |  |
| Negative symptoms | - | 15.6 ± 5.7 |  |
| Self-perceived functioning, n (%) |  |  |  |
| Excellent | - | 26 (17.4%) |  |
| Good | - | 49 (32.9%) |  |
| Fair | - | 61 (40.9%) |  |
| Poor | - | 13 (8.7%) |  |
| Earned income (No, %) |  | 111 (74.5%) |  |
| Planned activities, n (%) |  |  |  |
| Never_rare | - | 64 (42.7%) |  |
| Occasional | - | 32 (21.3%) |  |
| Regular | - | 54 (36%) |  |

Values are presented as mean ± SD, except for CPZ, which is presented as median (IQR). Categorical variables are presented as n (%). Sample sizes may vary slightly across variables because of missing data. HC: Healthy control; SCZ: Schizophrenia; CPZ: Total chlorpromazine equivalent dose; WTAR: Wechsler Test of Adult Reading; Processing speed: MCCB processing speed T-score; Attention: MCCB attention T-score; Working memory: MCCB working memory T-score; Reasoning: MCCB reasoning T-score; Planned activities: Frequency of planned activities with others; Employment: Employment in the past 6 months or currently. p < 0.05\*, p < 0.01\*\*

Compared with HC, patients with SCZ had fewer years of education and lower WTAR scores, as well as poorer performance across all MCCB cognitive domains (all p < 0.001). The SCZ group also had a lower proportion of females than the HC group (p = 0.006). No significant difference in age was observed between groups.

The NUSDAST cohort included 156 HC and 166 individuals with SCZ. Participants with SCZ had fewer years of education (12.2 ± 2.4 vs. 14.2 ± 2.8 years, p < 0.001) and a lower proportion of females (33.1% vs. 47.4%, p = 0.01) than HC, whereas no significant difference in age was observed between groups (34.9 ± 12.9 vs. 33.1 ± 14.1 years, p = 0.16).

### N-LCS and deviation metrics

Both parallel analysis and the MAP procedure supported a one-factor solution for N-LCS derived in the COBRE cohort. This solution showed high stability across bootstrap samples, with a mean Tucker congruence of 0.99 (95% CI 0.98–1.00), consistent with our previous report [8].

Compared with HC, patients with SCZ exhibited significantly lower CDM (p < 0.001, Cohen’s d = -1.60; Table 2), indicating a shared reduction across cognitive domains. CDA was centered around zero in HC but positively shifted in the SCZ group (p < 0.001, Cohen’s d = 1.51), suggesting notable alterations in the relationships among cognitive domains.

**Table 2.**
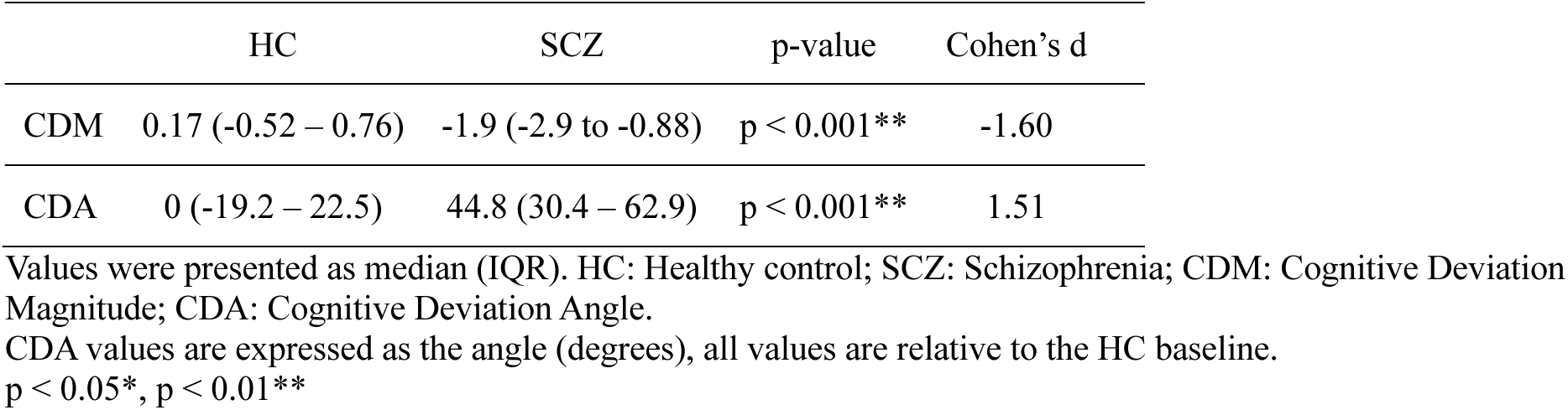
N-LCS deviation metrics in healthy controls and individuals with schizophrenia in COBRE cohort.

In the NUSDAST cohort, the one-factor structure of the N-LCS was also supported by parallel analysis and the MAP procedure. Bootstrap resampling demonstrated high stability of this solution, with a mean Tucker congruence of 0.99 (95% CI 0.99–1.00).

The N-LCS structures derived from the two cohorts showed high structural similarity, with a Tucker congruence of 0.90 and a cosine similarity of 0.89.

### N-LCS model development and internal validation

Model development and internal validation were performed using the nested bootstrap procedure, with performance estimates corrected for optimism. For OF, the final model retained CDM, age, education, and self-perceived functioning. The model achieved an AUC of 0.73 and balanced accuracy of 0.71. Calibration intercept and slope were 0.03 and 0.99, respectively (Table 3). Optimism-corrected DCA indicated net benefit across the full threshold range (0–0.99) for the OF model (Figure 1).

**Figure 1.**
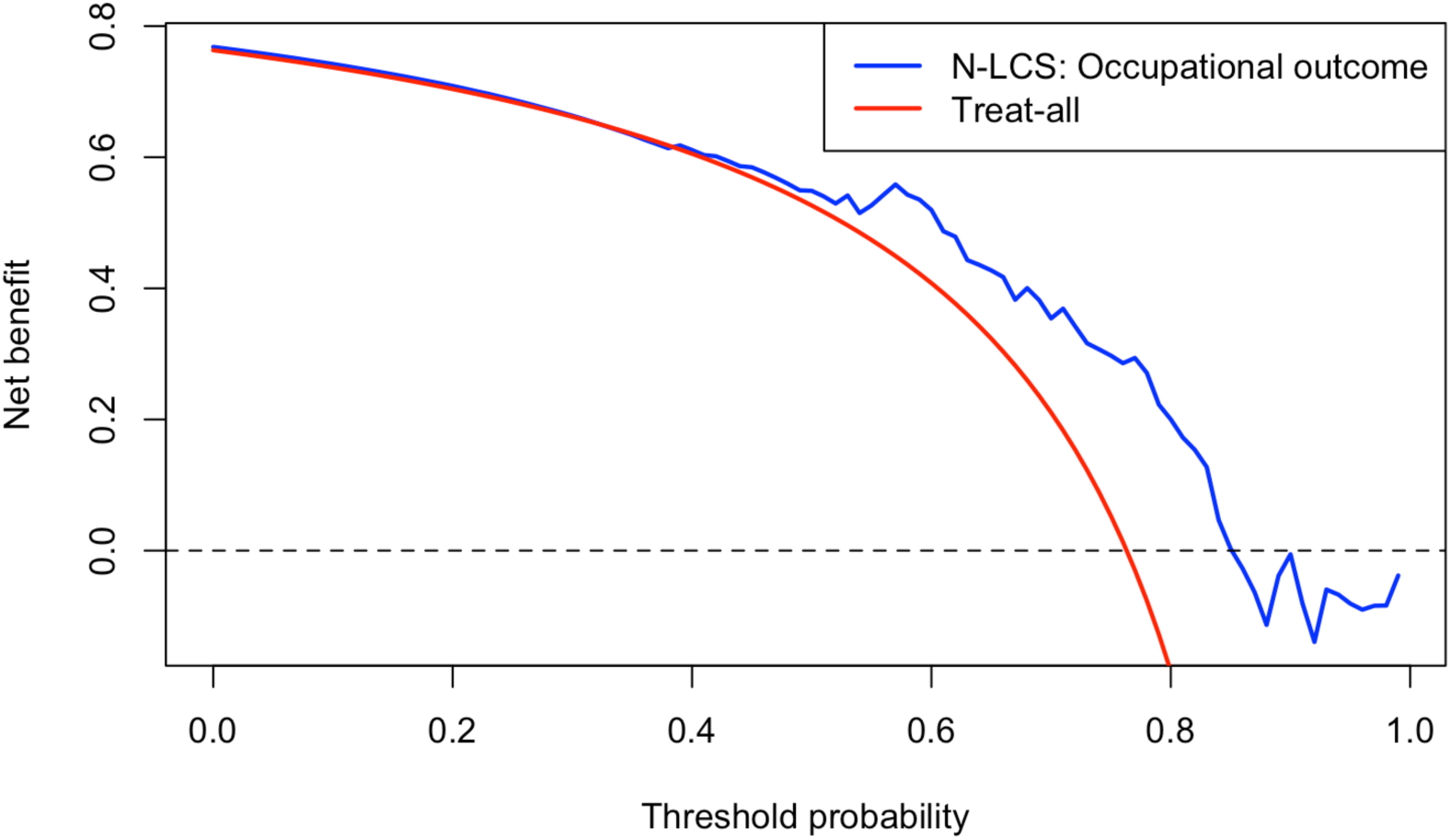
Decision curve analysis of occupational functioning outcomes

**Table 3.** Predictive performance of N-LCS-based and score-based models in COBRE cohort.

| Outcome | Occupational functioning |  |  | Social functioning |
| --- | --- | --- | --- | --- |
| Model | N-LCS (full model) | Score-based | N-LCS (reduced model) | N-LCS |
| N (events) |  | 114 (87) |  | 142 |
| Final predictors | 4 | 7 | 3 | 6 |
| EPV | 21.8 | 12.4 | 29 | - |
| Corrected AUC (Optimism) | 0.73 (0.06) | 0.75 (0.06) | 0.74 (0.03) | - |
| Correct Bal Acc (Optimism) | 0.71 (0.06) | 0.72 (0.07) | 0.73 (0.04) | - |
| Corrected sensitivity | 0.90 | 0.87 | 0.75 |  |
| Corrected specificity | 0.52 | 0.58 | 0.72 |  |
| Corrected calibration intercept | 0.03 | -0.09 | -0.61 | - |
| Corrected calibration slope | 0.99 | 1.06 | 1.54 | - |
| Corrected C-index (Optimism) | - | - |  | 0.66 (0.06) |
| Corrected weighted kappa (optimism) | - | - |  | 0.27 (0.09) |
| Corrected log-loss (optimism) | - | - |  | 1.03 (-0.05) |

N-LCS: Normative Latent Cognitive Structure; EPV: Events per variable; N (events): Total sample size used for the model, with No-income defined as the event in binary outcomes analyses; Correct Bal Acc (Optimism): Optimism-corrected balanced accuracy; optimism denotes bootstrap-estimated overfitting; AUC: Area under the receiver operating characteristic curve.

For SF, WTAR, negative symptoms, age, sex, education, and self-perceived functioning were retained in the final model. The C-index was 0.66, with a weighted kappa of 0.27 and log-loss of 1.03.

### External validation and recalibration of the N-LCS occupational functioning model

The OF-RM was externally validated in the NUSDAST cohort. The model demonstrated moderate discrimination, with an AUC of 0.70 (95% CI 0.58–0.79) and a balanced accuracy of 0.56 (95% CI 0.50–0.63). However, calibration was suboptimal, with a calibration intercept of -0.87 (95% CI -1.91 to 0.00) and a calibration slope of 0.48 (95% CI 0.20–0.85) (Table 4). DCA of the OF-RM showed positive net benefit across threshold probabilities of 0.15–0.99.

**Table 4.** External validation and recalibration of the N-LCS occupational functioning model.

| Outcome | Occupational functioning |  |
| --- | --- | --- |
| N (event) | 107 (67) |  |
| Number of predictors | 3 |  |
| EPV | 22.3 |  |
| Recalibration | Before recalibration | After recalibration |
| AUC | 0.70 (95% CI 0.58–0.79) | 0.70 (95% CI 0.58–0.79) |
| Bal Acc | 0.56 (95% CI 0.50–0.63) | 0.69 (95% CI 0.59–0.78) |
| Sensitivity | 0.97 (95% CI 0.92–1.00) | 0.82 (95% CI 0.72–0.90) |
| Specificity | 0.15 (95% CI 0.05–0.27) | 0.55 (95% CI 0.39–0.71) |
| Calibration intercept | -0.87 (95% CI -1.91 to 0.00) | - |
| Calibration slope | 0.48 (95% CI 0.20–0.85) | - |

N-LCS: Normative Latent Cognitive Structure; EPV: Events per variable; N (events): Total sample size used for the model, with Unemployed defined as the event in binary outcomes analyses; Bal Acc (Optimism): Optimism-corrected balanced accuracy; optimism denotes bootstrap-estimated overfitting; AUC: Area under the receiver operating characteristic curve.

After recalibration, discrimination remained unchanged (AUC 0.70, 95% CI 0.58–0.79), whereas balanced accuracy improved to 0.69 (95% CI 0.59–0.78).

### Comparison with score-based models

For OF, discrimination and balanced accuracy were comparable between the N-LCS and score-based models (Table 3). However, the N-LCS OF model showed superior optimism-corrected calibration, with both the calibration slope and intercept close to ideal. No score-based comparison was performed for SF, as the final model for this outcome did not retain a cognitive predictor.

## Discussion

This study developed and internally validated N-LCS prediction models for real-world functional outcomes in SCZ using a fully nested bootstrap procedure with explicit calibration and clinical utility evaluation, addressing important methodological limitations commonly reported in psychiatric prediction modeling [4]. The reduced OF model was further externally validated in an independent NUSDAST cohort, where recalibration improved model calibration and classification performance while preserving discrimination. To our knowledge, this is the first study to apply the N-LCS framework to clinical prediction, extending its application beyond characterizing cognitive heterogeneity to predicting real-world functional outcomes. Compared with conventional score-based cognitive representations, the N-LCS OF model showed comparable discrimination but better calibration, indicating that how cognition is represented may influence the agreement between predicted probabilities and observed outcomes.

The predictive utility of N-LCS depends on the reliable estimation of the underlying latent cognitive structure. In the present study, this one-factor N-LCS structure was replicated in both the COBRE and NUSDAST cohorts, supporting its stability across cohorts [33]. The high structural similarity between cohorts suggests that N-LCS may provide a common framework for characterizing cognitive function across studies using different assessment batteries, potentially improving the generalizability of prediction models that incorporate cognitive predictors across clinical settings.

The present study selected two objectively defined indicators spanning occupational and social domains of real-world functioning in SCZ. This selection reduces reliance on clinician- or self-rated impressions of overall functioning [22], and reflects concrete milestones relevant to patients’ daily lives [34]. However, there remains no consensus regarding the definition and operationalization of functional outcomes [35], and these indicators were collected via self-report interviews rather than independent verification [17] and thus remain subject to reporting inaccuracies despite their objective definition. Furthermore, real-world functioning is a multidimensional construct that may extend beyond the specific indicators examined here [36].

The candidate predictor pool was specified a priori based on prior evidence and their established use in prediction models, aiming to reduce selection bias and limit overfitting [37]. Other factors known to influence functional outcomes, including physical health, social support, and environmental factors, were unavailable in the present datasets and may provide additional predictive value in future studies [36].

Among the evaluated outcomes, the OF model demonstrated moderate-to-good predictive performance after optimism correction [38], with calibration approaching the ideal [31]. Several methodological features support the robustness of the OF model. Its EPV exceeded 20, surpassing the threshold advocated in recent statistical literature [30], and ridge regression was applied throughout model development in accordance with methodological recommendations for shrinkage [28], together mitigating the risk of overfitting and improving model stability. In addition, the use of a fully nested bootstrap procedure, incorporating predictor-set selection, penalty tuning, and threshold determination within the validation process, reduced optimism and satisfied current recommendations for rigorous internal validation [4]. Consistent with these methodological strengths, the favorable calibration of the OF model indicates that predicted probabilities were closely aligned with observed outcomes, supporting the reliability of its predictions [31].

Because self-perceived functioning was unavailable in NUSDAST, a reduced model (OF-RM) was used for external validation. It demonstrated discrimination comparable to that of the full model, indicating that CDM, age, and education retained most of the predictive information. Calibration was modestly reduced, suggesting that self-perceived functioning may have contributed additional information for more accurate individual risk estimation [31]. External validation in an independent cohort showed only a modest decrease in discrimination for the OF- RM, supporting its generalizability across cohorts. The calibration intercept shifted further from zero in NUSDAST, which may partly reflect the lower event rate compared with the COBRE cohort (63% vs. 76%). Balanced accuracy was lower when the original Youden index was applied but improved after recalibration, highlighting the importance of recalibration when prediction models are transported across clinical settings [39]. The generalizability of the full OF model, however, remains to be established.

The DCA results in the COBRE cohort provide preliminary evidence that the OF model offers net benefit over default strategies across nearly the entire range of threshold probabilities [40]. This wide range may be related to the high event rate observed in the analyzed sample. From a clinical perspective, the OF model represents a promising candidate for further evaluation toward clinical translation, supported by consistent findings across internal validation, external validation, and decision curve analysis. However, the moderate sample size, event imbalance, and need for further validation of the full model in larger independent cohorts limit conclusions regarding clinical implementation.

By contrast, the SF model demonstrated only modest predictive performance [23]. This may reflect differences in how these functional domains are operationalized as outcome variables and the extent to which each domain is captured by the prespecified predictors. The stable performance of the OF model across cohorts is consistent with the established relationship between occupational functioning and cognitive ability, with educational attainment and age contributing additional predictive information [41]. The retained predictor set for SF included negative symptoms but no cognitive predictor, consistent with evidence that cognition shows greater predictive value for vocational outcomes, whereas negative symptoms may contribute more to SF outcomes [42]. The remaining variability in SF may therefore be influenced by factors not captured in the present study, including interpersonal relationships and social support. Accordingly, the SF model should be considered preliminary and requires further refinement before external validation.

The N-LCS OF model achieved comparable predictive performance to the score-based model, with fewer predictors. One possible explanation is that N-LCS captures the shared latent cognitive structure across cognitive domains, thereby reducing redundancy while preserving predictive information [8]. From a clinical modeling perspective, achieving comparable predictive performance with fewer predictors may improve model parsimony, with the reduced predictor burden also resulting in a higher EPV [30]. The favorable calibration observed in the N-LCS model further supports greater reliability of predicted probabilities compared with score-based approaches [43]. This may be explained by a closer correspondence between the structure-informed cognitive representation and functional outcomes, leading to probability estimates that more closely align with observed outcome frequencies.

Several limitations should be considered. First, given the moderate sample size of the present study, replication in larger independent cohorts with different clinical characteristics is required to establish the generalizability and robustness of these findings. Second, the cross-sectional design precluded evaluation of whether N-LCS predicts longitudinal trajectories of functional outcomes. Third, because ridge regression was applied to reduce overfitting through coefficient shrinkage, predictor-level inference was not an objective of the present analysis. The contribution of individual predictors therefore warrants further investigation using methods better suited to predictor interpretation.

In conclusion, this study demonstrates the potential of N-LCS as a structure-informed cognitive representation for prediction modeling in SCZ. Among the evaluated outcomes, the OF model showed the most favorable predictive performance, supported by good calibration, preliminary external validation, and DCA evidence, whereas the SF model remains at a preliminary stage requiring further predictor development. These findings support continued evaluation of the OF model in larger independent cohorts to further establish its generalizability and potential clinical utility.

## Data Availability

The datasets analyzed in this study are publicly available through the Collaborative Informatics and Neuroimaging Suite Data Exchange tool (COINS; http://coins.mrn.org/dx) and the Northwestern University Schizophrenia Data and Software Tool (NUSDAST; https://central.xnat.org/).

## Acknowledgements

Data were downloaded from the Collaborative Informatics and Neuroimaging Suite Data Exchange tool (COINS; http://coins.mrn.org/dx) and data collection was performed at the Mind Research Network and funded by a Center of Biomedical Research Excellence (COBRE) grant 5P20RR021938/P20GM103472 from the NIH to Dr. Vince Calhoun. Additional data collection was funded by NIMH R01MH084898-01A1, “Brain Glutamate and Outcome in Schizophrenia”, PI: J. Bustillo. Data for external validation were obtained from the Northwestern University Schizophrenia Data and Software Tool (NUSDAST) and downloaded from XNAT Central (https://central.xnat.org/).

The investigators associated with these datasets contributed to the design of the original research and data collection but did not participate in the analyses reported in this manuscript or in the preparation of this manuscript. The author gratefully thanks them for making these datasets publicly available.

## Funding

This work was not supported by any specific funding.

## Declaration of Interest

The author declares no conflicts of interest.

## Notes

### Competing Interest Statement

The authors have declared no competing interest.

### Author Declarations

The datasets analyzed in this study are publicly available through the Collaborative Informatics and Neuroimaging Suite Data Exchange tool (COINS; http://coins.mrn.org/dx) and the Northwestern University Schizophrenia Data and Software Tool (NUSDAST; https://central.xnat.org/). All individual-level data were de-identified prior to use in this study.

### Summary of Updates

Added external validation using an independent schizophrenia cohort and demonstrated the structural similarity of the Normative Latent Cognitive Structure (N-LCS) across cohorts with different cognitive assessment batteries.

